# Antibody-mediated protection against symptomatic COVID-19 can be achieved at low serum neutralizing titers

**DOI:** 10.1101/2022.10.18.22281172

**Authors:** Pete Schmidt, Kristin Narayan, Yong Li, Chengzi I. Kaku, Michael E. Brown, Elizabeth Champney, James C. Geoghegan, Maximiliano Vásquez, Eric M. Krauland, Thomas Yockachonis, Shuangyi Bai, Bronwyn M. Gunn, Anthony Cammarata, Christopher M. Rubino, Laura M. Walker

**Affiliations:** Invivyd, Inc., Waltham, MA 02451, USA; Adimab LLC, Lebanon, NH 03766, USA; Paul G. Allen School of Global Health, Washington State University, Pullman, WA 99164, USA; Institute for Clinical Pharmacodynamics, Schenectady, NY 12305

**Author notes:** Corresponding author. (L.M.W.).

## Abstract

Multiple studies of vaccinated and convalescent cohorts have demonstrated that serum neutralizing antibody (nAb) titers correlate with protection against COVID-19. However, the induction of multiple layers of immunity following SARS-CoV-2 exposure has complicated the establishment of nAbs as a mechanistic correlate of protection (CoP) and hindered the definition of a protective nAb threshold. Here, we show that a half-life extended monoclonal antibody (adintrevimab) provides approximately 50% protection against symptomatic COVID-19 in SARS-CoV-2-naïve adults at low serum nAb titers on the order of 1:30. Vaccine modeling supports a similar 50% protective nAb threshold, suggesting low levels of serum nAb can protect in both monoclonal and polyclonal settings. Extrapolation of adintrevimab pharmacokinetic data suggests that protection against susceptible variants could be maintained for approximately 3 years. The results provide a benchmark for the selection of next-generation vaccine candidates and support the use of broad, long-acting monoclonal antibodies as an alternative or supplement to vaccination in high-risk populations.

---

The establishment of correlates of protection (CoPs)−defined as specific immunological markers associated with protection against infection or disease caused by a pathogen (*1*)−is of critical importance for accelerating the licensure of new vaccines, evaluating susceptibility to disease, and determining the need for and optimal timing of booster vaccinations. There are two types of CoPs: mechanistic CoPs, which are directly responsible and statistically interrelated with protection, and non-mechanistic CoPs, which are correlated with the mechanistic factor without directly conferring protection (*1*).

Multiple studies in both humans and animal models have demonstrated that the induction of nAb responses correlates with protection against COVID-19 (*2-8*). However, all of the human CoP studies performed to date have been confounded by the presence of other forms of immunity induced by natural infection and vaccination (e.g., memory B cells and T cells). These other immune responses may also contribute to, and independently correlate with, protection and therefore complicate the establishment of serum nAb as a mechanistic correlate of protection. Furthermore, the lack of standardized serological assays has required the normalization of vaccine-induced nAb titers to those observed following convalescence, allowing only for the determination of relative, rather than absolute, protective titers (*3, 8*). Finally, although clinical studies have demonstrated that passively transferred monoclonal antibodies can protect against COVID-19 in the absence of other forms of immunity, the high levels of serum neutralization conferred by these therapies has precluded the definition of a protective neutralization threshold (*9, 10*).

We conducted a Phase 2/3 clinical study to evaluate the efficacy of a half-life extended human monoclonal antibody (adintrevimab) in the prevention of symptomatic COVID-19 in SARS-CoV-2-naïve adults during the emergence and global spread of SARS-CoV-2 variants Delta and Omicron BA.1/BA1.1. Due to marked differences in adintrevimab potency against these two variants (*11*), combined with the natural waning of passively transferred nAb titers over time, we were afforded the unique opportunity to assess the relationship between neutralization titer and clinical protection against symptomatic COVID-19 in the absence of pre-existing SARS-CoV-2 immunity. The observed protective efficacy of adintrevimab in our trial, combined with vaccine modeling data, provide strong evidence that nAbs are mechanistic in mediating protection against symptomatic COVID-19 and suggest that clinically meaningful efficacy can be achieved at low serum neutralizing titers on the order of 1:30.

## Results

We previously described a human IgG1 broadly neutralizing antibody (bnAb), ADG2, directed to the SARS-CoV-2 receptor binding domain (*12*). The precursor to this bnAb was isolated from a 2003 SARS survivor and engineered in vitro to improve its neutralization breadth and potency against a wide range of hACE2-using sarbecoviruses (*12, 13*). We introduced a two amino acid modification (M428L/N434A; “LA”) into the Fc region of ADG2 (ADG2-LA; hereafter referred to as adintrevimab) to enhance binding affinity to the neonatal Fc receptor under the acidic conditions of the lysosome (pH 6.0) and prolong serum half-life in vivo. Previous studies have shown that these two individual mutations improve binding to human FcRn at low pH, and the N484A variant has been shown to exhibit significantly decreased clearance relative to wild-type in rhesus macaques (*14-16*). In accordance with these studies, adintrevimab bound with 6-fold and 5-fold higher affinity to human and cynomolgus macaque FcRn, respectively, at pH 6.0 relative to ADG2 (Fig. 1A).

**Figure 1.**
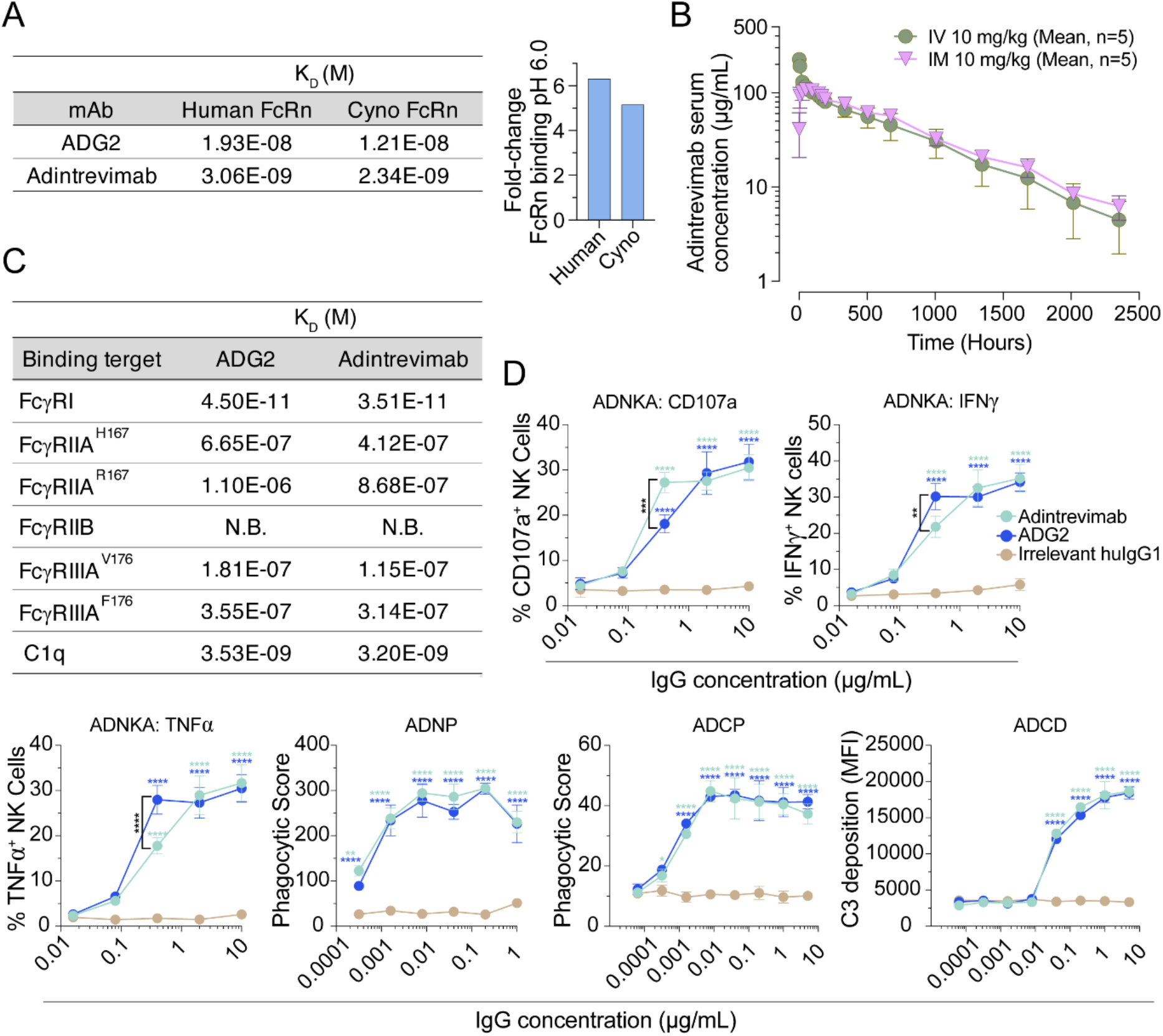
Adintrevimab shows intact Fc-mediated effector functions and extended half-life in NHPs. (A) Binding affinities of ADG2 and adintrevimab to human and cyno FcRn at pH 6.0, as measured by surface plasmon resonance (SPR). (B) Mean serum concentrations of adintrevimab in cynomolgus monkeys following administration of a single 10 mg/kg i.v. or i.m. dose. Error bars represent the standard deviations. (C) Binding affinities of ADG2 and adintrevimab to human FcγRs, as measured by SPR, and C1q, as determine by biolayer interferometry. (D) In vitro Fc-mediated functional activities of ADG2 and adintrevimab. Primary human NK cells were analyzed for surface expression of CD107a, indicating degranulation (top row, left panel), and the production of interferon-γ (IFNγ) (top row, right panel) or tumor necrosis factor–α (TNFα) (bottom row, first panel) following incubation with antibody-RBD immune complexes. Antibody-mediated phagocytosis of RBD-coated beads by differentiated THP-1 monocytes (bottom row, second panel) or HL-60 neutrophils (bottom row, third panel). Antibody-mediated complement deposition was measured by detection of complement component C3 onto RBD-coated beads after incubation of guinea pig complement with immune complexes (bottom row, fourth panel). Data points and error bars represent mean and SD, respectively. All data are representative of two independent experiments. Statistical comparisons were determined by two-way ANOVA with Tukey’s multiple comparison test. MFI, mean fluorescence intensity; N.B., non-binding; \**P* < 0.05, \*\**P* < 0.01, \*\*\**P* < 0.001, \*\*\*\**P* < 0.0001.

To determine whether the improvement in FcRn binding at low pH translated to extended serum half-life in vivo, we evaluated the PK of adintrevimab for up to 98 days in naïve cynomolgus macaques following a single 10 mg/kg IM injection or IV infusion. Following IV infusion, we observed low clearance (mean = 0.126 ml/h/kg) and a mean half-life (T_1/2_) of 19.7 days compared with 8-10 days for non-Fc modified IgGs in NHPs (*17*) (Fig. 1B). Systemic exposure following a single IM dose at 10 mg/kg demonstrated high relative bioavailability (111%) with similarly long half-life (T1/2 mean = 22.2 days) (Fig. 1B and Table S1). Thus, the LA modification endowed adintrevimab with an extended half-life in NHPs.

Because mutations that enhance binding to FcRn can adversely affect binding to human Fcγ receptors and the complement component C1q (*18*), we evaluated the impact of the LA modification on binding to recombinant FcγRI, FcγRIIa, FcγRIIb, FcγRIIIa, and C1q. We also assessed antibody-dependent natural killer cell activation and degranulation (ADNKA), antibody-dependent cellular phagocytosis mediated by monocytes and neutrophils (ADCP and ADNP), and antibody-mediated complement deposition (ADCD) using in vitro effector function assays. ADG2 and adintrevimab displayed comparable binding affinities to human Fcγ receptors and C1q and induced similar levels of ADNKA, ADCP, ADNP, and ADCD activity in vitro, suggesting that the LA modification enhances FcRn binding without substantially impacting Fc-mediated effector activities (Fig. 1C, D). Finally, we confirmed that the LA modification did not significantly affect the biophysical properties of the molecule, as assessed by in vitro polyreactivity, hydrophobicity, and thermal stability assays (*19*) (Fig. S1).

Based on its neutralization profile, biophysical properties, and extended half-life in NHPs, we advanced adintrevimab into a Phase 1 study to assess safety and pharmacokinetics and a Phase 2/3 prevention study with a pre-exposure prophylaxis (PrEP) cohort (EVADE; NCT04859517) to evaluate the ability of a single 300 mg dose to prevent the development of symptomatic COVID-19 in SARS-CoV-2-naïve adults. In the Phase 2/3 study, the primary analysis population comprised all participants who were seronegative and RT-PCR negative at baseline. The primary efficacy endpoint was symptomatic COVID-19 (SARS-CoV-2 infection confirmed by reverse-transcriptase–polymerase-chain-reaction assay) occurring after administration of adintrevimab or placebo through day 90. The study enrolled between 27 Apr 2021 and 11 January 2022, which spanned the transition from Delta to Omicron BA.1/BA1.1as the dominant circulating variant (Fig. S2).

Given the significant (>100-fold) loss of adintrevimab neutralizing activity against Omicron BA.1/BA1.1 relative to the Delta variant (Fig. S3) (*11*), which we hypothesized would translate into a substantial reduction in clinical efficacy, the trial population was divided into two subsets for the purposes of this analysis: the “pre-Omicron population”, which consisted of participants enrolled on or prior to 30 November 2021 and with events on or prior to 15 December 2021 (the date Omicron BA.1 became the predominant circulating variant in study locations), and the “Omicron population”, which consisted of participants enrolled between 01 December 2021 and 11 January 2022 and with events occurring prior to 11 April 2022. Whole-genome sequencing (WGS) on a subset of trial participants confirmed that the vast majority (97.7%) of infections in the pre-Omicron population were caused by the Delta variant, and 90.5% of infections in the Omicron population were caused by Omicron BA.1 or BA1.1 variants (Fig. S2 and table S2).

In the pre-Omicron population, adintrevimab demonstrated a relative risk reduction of 71% versus placebo in the development of RT-PCR confirmed symptomatic COVID-19 through 3 months, the primary endpoint. A post-hoc analysis of a subset of pre-Omicron EVADE participants randomized prior to 15 Jun 2021 revealed an 84% relative risk reduction vs placebo in the development of RT-PCR confirmed symptomatic COVID-19 through 6 months (Table S3). The increased efficacy through day 180 was driven by a greater percentage of events in the placebo group than in the adintrevimab group during months 3 through 6, as compared with months 0 through 3. Efficacy waned more rapidly in the Omicron PrEP population, as expected given the lower potency of adintrevimab against this variant. Here we observed a relative risk reduction of 60%, 41%, and 37% versus placebo in the development of RT-PCR confirmed symptomatic COVID-19 through day 28, 60, and 90, respectively (Table S3).

The dramatic difference in serum neutralizing activity conferred by adintrevimab against Delta and Omicron BA.1/BA1.1 provided us with the opportunity to potentially determine a threshold level of neutralization associated with protection against symptomatic COVID-19 in the absence of pre-existing immunity. As participants in the primary and exploratory efficacy analysis populations were primarily infected with Delta and Omicron BA.1/BA1.1, respectively, we normalized our population PK model-derived median serum concentrations of adintrevimab to authentic virus neutralization data for Delta and Omicron BA.1/BA1.1 to project serum nAb titers against these variants over time (serum neutralization titer = adintrevimab serum concentration/variant IC_50_) (*11*) (Table S4 and Fig. 2A). Based on this analysis, adintrevimab-mediated serum neutralization against the Delta variant peaked on day 8 with a median neutralizing titer of 1:6157 and declined to a median titer of 1:987 on day 360 (Fig. 2A). Because adintrevimab neutralizes Omicron BA.1 and BA1.1 with approximately 180-fold lower potency than Delta (Fig. S3) (*11*), the projected serum neutralizing titers peaked at proportionally lower levels (1:34 on day 8) against Omicron BA.1/BA1.1 and declined to <1:20 titers by day 120 (Fig. 2A). To validate the calculation used for converting serum concentrations to neutralizing titers, we also experimentally measured authentic virus serum neutralizing titers in all participants in our Phase 1 study, which showed that the normalized and measured serum neutralizing titers were within 3-fold for both Delta and Omicron BA.1 (Fig. S4). For comparison, we also measured vaccine-induced serum neutralizing titers against Delta and Omicron BA.1 in 12 healthy adult donors who had received a second dose of an mRNA vaccine (mRNA-1273) 7- to 30-days prior to sampling (Table S5). In this cohort, serum neutralizing titers ranged from 1:260-1:1024 with a geometric mean titer of 1:479 against the Delta variant (Fig. 2B). Consistent with prior studies (*11*), we observed significantly lower vaccine-induced serum neutralizing titers against the Omicron BA.1 variant, which ranged from <1:24-1:53 with a geometric mean titer of 1:31 (Fig. 2B). We conclude that administration of a 300 mg dose of adintrevimab results in high serum neutralizing titers of >1:500 against the Delta variant for at least one year, with geometric mean titers at 6 months exceeding peak titers achieved following two doses of an mRNA vaccine, whereas the same dosing regimen results in low serum neutralizing titers against Omicron BA.1 even at peak concentrations.

**Figure 2.**
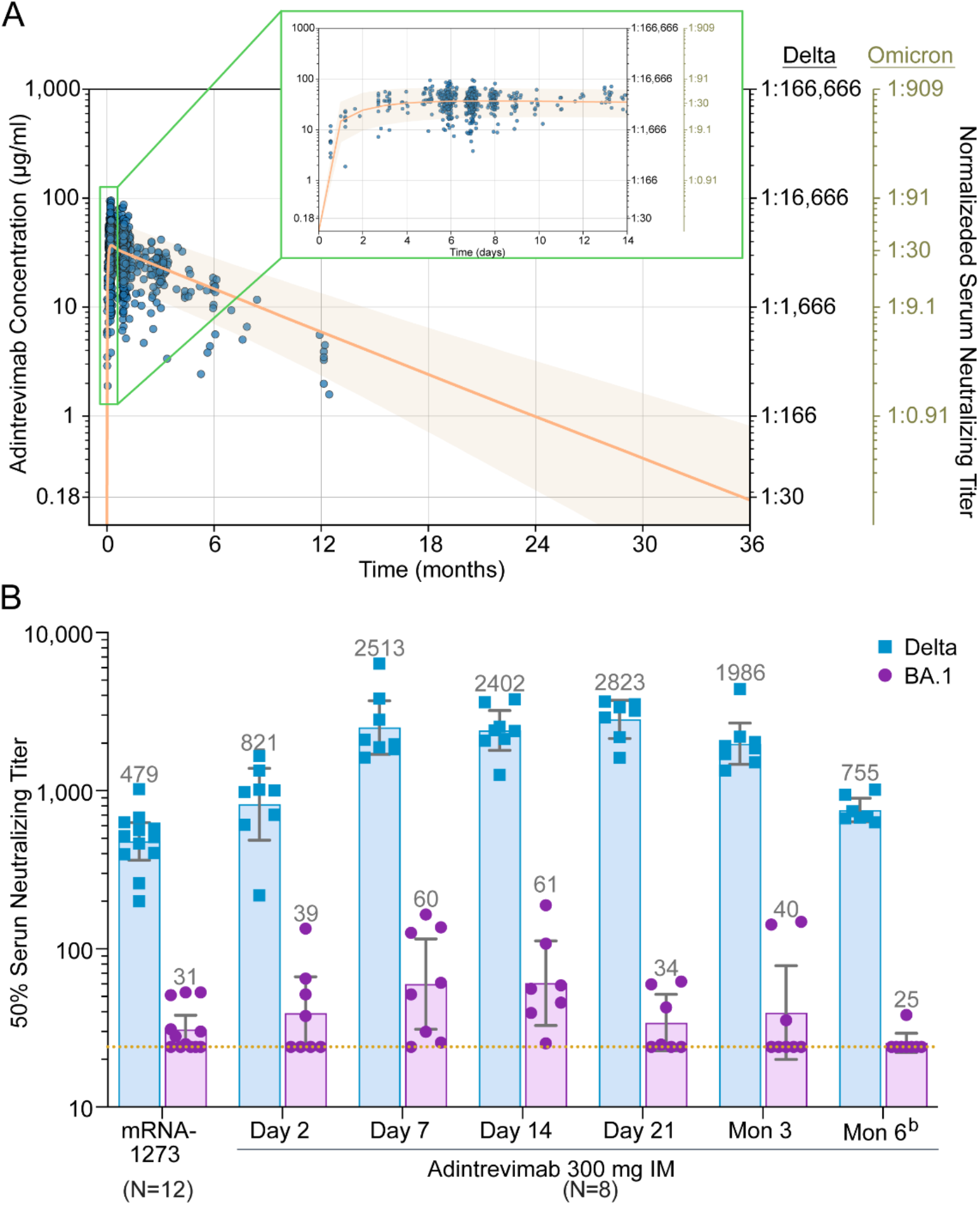
Adintrevimab pharmacokinetic parameters and neutralizing titers in humans following a single 300 mg IM dose. (A) Two-compartment population PK predicted adintrevimab serum concentrations and normalized Delta and Omicron BA.1 serum neutralizing titers at the indicated time points. Data points represent observed concentration-time data for subjects enrolled in Phase 1, EVADE (NCT04859517), and STAMP (NCT04805671). The solid line indicates median predicted concentration over time, and the shaded region shows the 5th to 95th percentiles of the predicted concentrations from the model-based simulations. (B) Measured geometric mean serum nAb titers against Delta and Omicron BA.1 at the indicated time points following a single 300 mg intramuscular injection of adintrevimab in healthy adult participants (*n* = 8) or 7-30 days following the second dose of mRNA-1273 (*n* = 12). Bar heights and error bars represent geometric mean FRNT_50_ titer ± SD, and the geometric mean PRNT_50_ titer is indicated above each bar. The dotted line represents the limit of detection for Omicron BA.1 titers based on a starting serum dilution of 1:24. Results from samples with BA.1 titers <24 were imputed to 24 in order to calculate geometric mean titers. Data were excluded from phase 1 study participant samples following confirmed COVID-19 infection or vaccination. Individuals receiving mRNA-1273 had no history of prior SARS-CoV-2 infection or vaccination.

We next investigated the relationship between adintrevimab-conferred serum neutralizing titers and protective efficacy observed in our prevention trial. On days 90 and 180, the normalized Delta serum neutralizing titers were 1:3880 and 1:2454, respectively, which was associated with 71% (day 90) and 84.4% (day 180) risk reductions in the development of symptomatic Delta infection (Fig. 3A and Table S3). On days 28, 60, and 90, the median serum concentrations of adintrevimab were 31.2, 27.1, and 23.28 mg/L, respectively, which translated to serum neutralizing titers of 1:29, 1:25, and 1:21, respectively against Omicron BA.1. These neutralizing titers corresponded to 60% (day 28), 41% (day 60), and 37% (day 90) relative risk reductions in the development of symptomatic Omicron BA.1/BA1.1 infection, thus demonstrating that adintrevimab provided measurable protection against symptomatic COVID-19 even at serum nAb titers as low as approximately 1:30 (Fig. 3A and Table S3).

**Figure 3.**
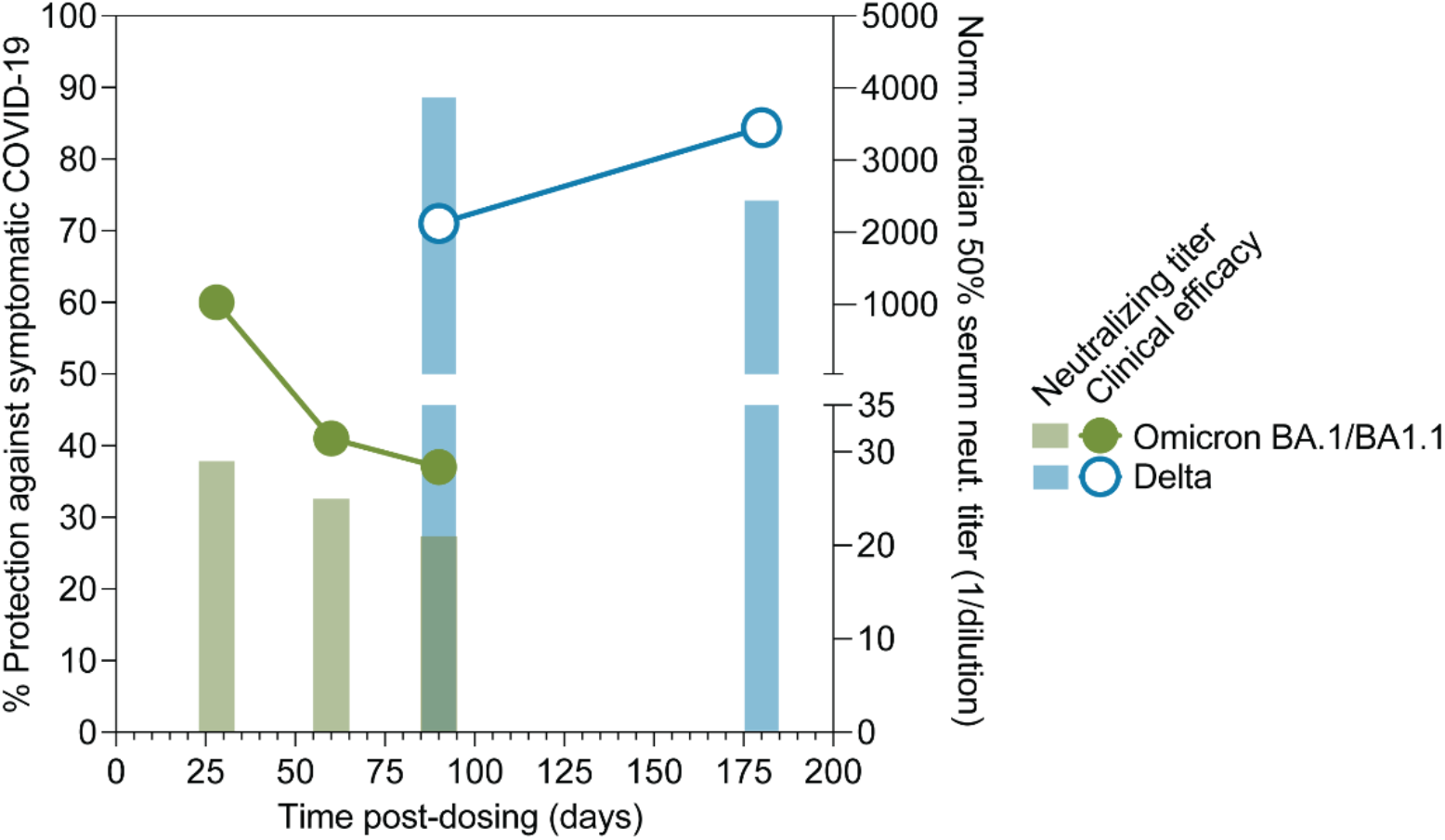
Serum neutralizing titers and clinical efficacy of adintrevimab against Delta and Omicron BA.1/BA1.1 variants. (A) Blue and green symbols indicate observed adintrevimab efficacy against symptomatic Delta or Omicron BA.1/BA1.1 infection, respectively, through the indicated time points following administration of a 300 mg dose. Solid blue and green bars indicate normalized adintrevimab serum neutralizing titers against Delta and Omicron BA.1/BA1.1, respectively, at the indicated time points. Normalized serum neutralizing titers were calculated by dividing median adintrevimab serum concentration by the variant IC_50._ Norm., normalized; Neut., neutralization.

Passive transfer experiments in animal models have demonstrated that certain anti-viral antibodies are unusually potent in protection relative to their neutralization abilities (*7, 20*). To determine whether adintrevimab protects at lower serum neutralizing titers than polyclonal nAbs induced by vaccination, we investigated the relationship between geometric mean serum neutralizing titers induced following ChAdOx1 or bNT162b2 vaccination−as measured in the same authentic virus FRNT assay used to determine normalized adintrevimab serum neutralizing titers (*11, 21, 22*)−and protection against symptomatic COVID-19 reported in phase 3 clinical trials and/or real-world vaccine effectiveness studies (*23-37*) (Table S6). Despite uncontrolled variables across the vaccine studies, we observed a strong correlation between geometric mean FRNT_50_ titers and protection against symptomatic COVID-19 across different vaccine platforms, SARS-CoV-2 variants, and time intervals post-vaccination (Fig. S5). Fitting the data to a nonlinear regression model allowed us to determine a 50% protective serum neutralization titer of 1:31 (95% confidence interval [CI], 1:24-1:40) (Fig. S5 and Fig. S6). However, as observed in prior vaccine modeling studies (*3, 8*), the shape of the protection curve suggests that protection increases gradually with neutralization titer, with no absolute threshold above which protection is achieved.

To test the predictive utility of the model, we used normalized neutralizing titers for adintrevimab to predict clinical efficacy against Delta and Omicron BA.1/BA1.1. The model predicted an efficacy of 92.8% for adintrevimab against Delta on day 180, which was similar to the observed efficacy in our prevention trial, where individuals in the adintrevimab arm experienced an 84.4% relative risk reduction in the development of symptomatic disease through 6 months (Fig. S6). Similarly, the model predicted an efficacy of 47.7% (95% CI, 39.9%– 55.4%), 42.5% (95% CI, 34.4%–50.6%), and 37.2% (95% CI, 28.9%–45.4%) against Omicron BA.1 on days 28, 60, and 90, respectively. These predicted efficacies were within the range of the observed efficacies of 60%, 41%, and 37% at a median follow-up of 28, 60, and 90 days, respectively (Fig. S6). Inclusion of the observed adintrevimab efficacy data in the vaccine CoP model resulted in a comparable 50% protective neutralizing titer of 1:30 (95% CI, 1:25-1:35) (Fig. 4A and Fig. S6). Notably, extrapolation of serum adintrevimab concentration over time demonstrated that adintrevimab-conferred serum neutralizing titers against the Delta variant would remain over this 50% protective neutralization titer for approximately 3 years (Fig. 2A), suggesting that potent, half-life extended monoclonal antibodies have the potential to provide prolonged protection against susceptible SARS-CoV-2 variants.

**Figure 4.**
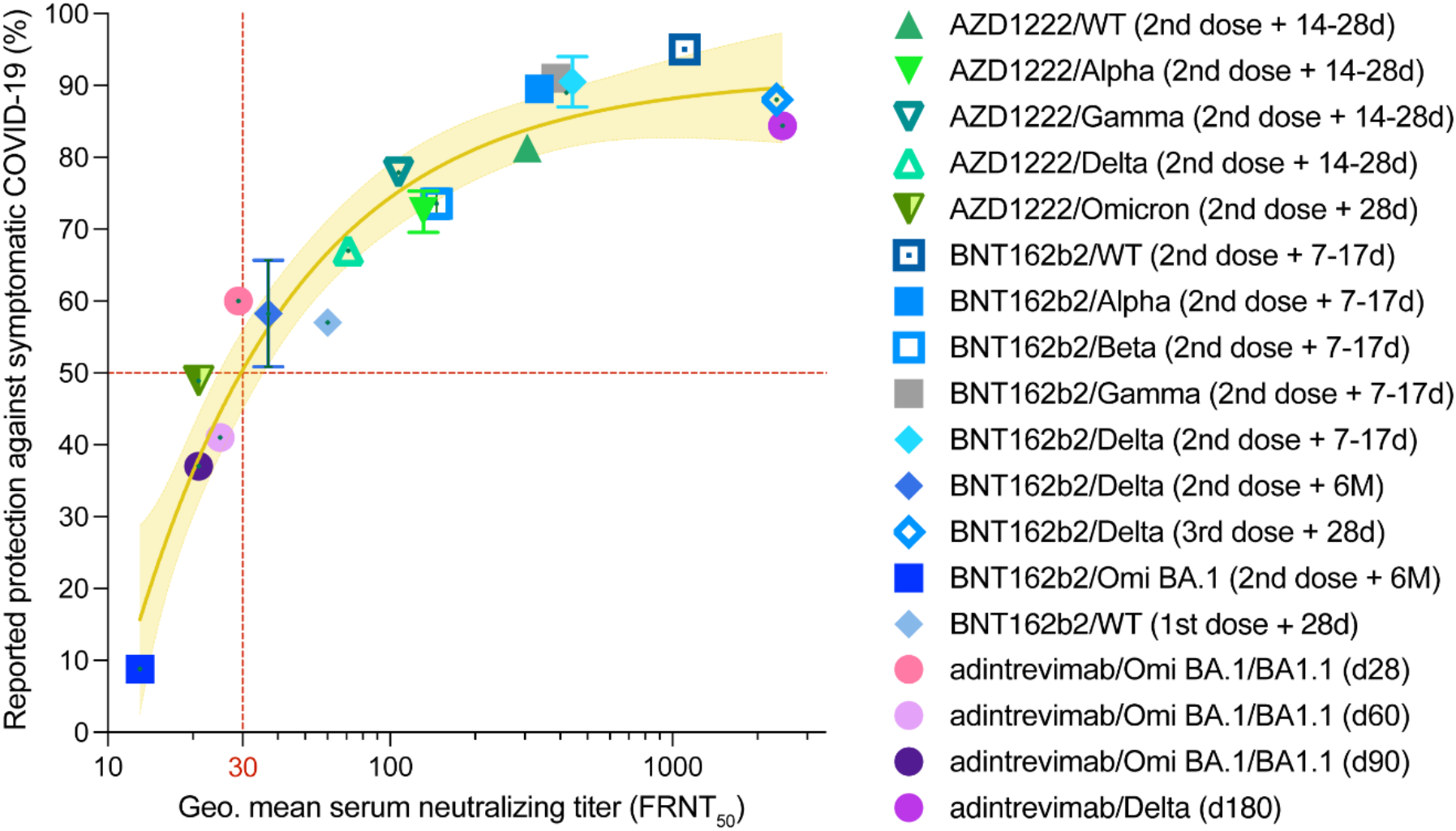
Relationship between median serum neutralization titer and protective efficacy against symptomatic COVID-19. (A) Reported median vaccine-induced serum neutralizing titers and normalized monoclonal antibody neutralizing titers (adintrevimab), measured in an authentic virus neutralization assay (*11, 21, 22*), plotted against reported efficacy in Ph3 clinical trials and/or real-word vaccine effectiveness studies (*23-37*). The number of vaccine doses and timing of blood sample collection and corresponding vaccine effectiveness or monoclonal antibody study read-outs against indicated variants are shown in parenthesis in the legend. The brown solid line indicates the best fit of the non-linear regression and the yellow shading indicates 95% confidence intervals. Data points and error bars represent mean ± SD. The neutralizing titer associated with 50% protection against symptomatic COVID-19 is indicated by a vertical dotted red line. Omi, Omicron; Geo., geometric.

In conclusion, our study demonstrates that both monoclonal antibody- and vaccine-mediated protection against symptomatic COVID-19 can be achieved at low serum nAb titers on the order of 1:30. However, it is important to note that this titer is not absolute, as breakthrough infections still occurred at timepoints associated with very high levels of serum neutralization. Similar results have been observed in the context of respiratory syncytial virus, where the probability of infection decreased with increasing levels of antibody, but breakthrough infections still occurred at high antibody titers suggesting that an absolute threshold of protection does not exist (*38*). The reasons for this are not well understood but may relate to individual differences in viral exposure factors (e.g. varying levels of virus in transmitting inoculums) or variability in antibody transudation into mucosal surfaces. Based on the observed efficacy of adintrevimab, as well as the reported efficacies of AZD7442 and REGEN-COV in a PrEP setting (*9, 10*), it appears that the maximum level of protection that can be achieved with neutralizing antibody alone is about 85%. In contrast, certain mRNA vaccines have demonstrated peak efficacies of about 95%, despite inducing similar or lower levels of serum neutralization as those achieved in monoclonal antibody efficacy studies (*27, 30*). Thus, although the majority of vaccine-mediated efficacy against symptomatic disease appears to be driven by nAbs, other mechanisms such as T cell and memory B cell responses, non-neutralizing antibodies, and/or innate immunity likely also contribute to protection and is it possible that their contribution increases in the setting of sub-protective antibody titers (*6*). Nevertheless, the 50% protective neutralizing titer defined here could be used to guide the advancement of next-generation vaccine candidates and monoclonal antibody dosing levels and intervals.

The high level of efficacy conferred by adintrevimab against symptomatic Delta infection through 6 months, combined with PK modeling showing maintenance of neutralizing titers associated with 50% protection against symptomatic disease for about 3 years, suggests that broad, highly potent, and half-life extended monoclonal antibodies have the potential to offer more durable protection than currently available vaccines. This advantage may be especially dramatic in the context of antigenically divergent variants, such as Omicron, given the limited magnitude and duration of protection conferred by vaccination. However, it is well-established that the rapid evolution of SARS-CoV-2 has also stymied therapeutic monoclonal antibodies, with the majority of clinical-stage and EUA authorized antibodies showing little to no neutralizing activity against Omicron BA.1 and/or its sublineages (*39, 40*). Thus, the future challenge will be in the development of next-generation antibody therapeutics that recognize functionally constrained and antigenically invariant epitopes, such as the highly conserved S2 stem helix region (*41-43*), and/or in modifying the current regulatory paradigm such that potent but relatively narrow-spectrum RBD antibodies can be developed and deployed at a pace that keeps up with viral evolution.

## Data Availability

Antibody sequences have been deposited in GenBank under accession codes MZ439266-MZ439267. IgGs are available from the corresponding author under MTA from Invivyd, Inc. All other data is available in the manuscript or supporting material.

## Acknowledgements

We thank A. Wec and B. West for assistance with figure preparation and Amanda Copans and Pamela Hawn for helpful comments on the manuscript. All of the antibody developability studies were performed by Adimab’s protein analytics group. We thank the EVADE study team for support of this project. We thank the team at WuXi AppTec for conducting the NHP PK study, and the teams at Q2 Solutions, Viroclinics, and Viracor (including T. Battle, E. Morris, J. Abbott, T. Paprotka, T. Brefort, M.Schmitt, Y. Kumar, J. Nutt, S. Cowden, K. Murray, M. Diaz, M. Watters, C. Marti, E. Bixler, T. Moss, and R. Sinha) for clinical sample analysis.

## Author contributions

L.M.W. and P. S. conceived and designed the study. M.E.B., E.C., and J.C.G. designed and supervised binding and developability and biolayer interferometry assays. K.N. and C.I.K. designed and analyzed results from serum neutralization assays. C.M.R., Y.L., and A.C performed pharmacokinetic modeling studies. M.V. and E.M.K. aided in engineering of adintrevimab and supervised parts of the in vitro preclinical characterization. T.J.Y, S.B., and B.M.G. designed and performed Fc-effector functional assays. C.I.K., K.N., P.S., and L.M.W. analyzed the data. P.S. and L.M.W. wrote the manuscript and all authors reviewed and edited the paper.

## Supplementary Materials

Materials and Methods

Figures S1-S6

Tables S1-S6

References (44-48)

