## Supplementary materials for "Antibody-mediated protection against symptomatic COVID-19 can be achieved at low serum neutralizing titers"

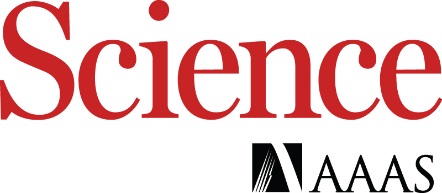

Supplementary Materials for

**Antibody-mediated protection against symptomatic COVID-19 can be achieved at low serum neutralizing titers**

Pete Schmidt^1^, Kristin Narayan^1^, Yong Li^1^, Chengzi I. Kaku^2^, Michael E. Brown^2^, Elizabeth Champney^2^, James C. Geoghegan^2^, Maximiliano Vásquez^2^, Eric M. Krauland^2^, Thomas Yockachonis^3^, Shuangyi Bai^3^, Bronwyn M. Gunn^3^, Anthony Cammarata^4^, Christopher M. Rubino^4^, Laura M. Walker^1^*

**This PDF file includes:**

Materials and Methods

Supplementary Figure Text

Figs. S1 to S6

Tables S1 to S6

**Materials and methods**

### Phase 1 clinical study design

ADG-001-01 is a Phase 1, randomized, double-blind, single-ascending-dose study to evaluate the safety and tolerability, pharmacokinetics, and immunogenicity of a single intramuscular (IM) or intravenous (IV) dose of adintrevimab or placebo administered to healthy participants while confined to the clinical unit. A total of 60 participants were enrolled in the study across 6 cohorts. In each cohort, 10 participants were randomized to receive adintrevimab (8 participants) or placebo (2 participants). Ex vivo serum neutralizing activity of adintrevimab against SARS-CoV-2 was also assessed.

**EVADE clinical study design**

EVADE is a Phase 2/3, multicenter, double blind, placebo controlled, randomized study of the mAb adintrevimab in the prevention of symptomatic COVID-19 in adults and adolescents with no known history of SARS-CoV-2 infection but whose circumstances place them at increased risk of acquiring SARS-CoV-2 infection and developing symptomatic COVID-19. This objective was independently evaluated in a cohort of participants with reported recent exposure to an individual diagnosed with a SARS-CoV-2 infection (Cohort A; post-exposure prophylaxis) and in a cohort of participants with no reported exposure to SARS-CoV-2 (Cohort B; pre-exposure prophylaxis). These cohorts were enriched for participants whose advanced age (≥55 years old) or health status places them at risk for severe COVID-19 or COVID-19 complications.

**Trial participants**

The pre-exposure prophylaxis (PrEP) cohort consists of adolescents (12-17 years of age) and adults ≥18 years of age who had an increased risk of exposure to SARS-CoV-2 owing to vocational or social circumstances. All participants were required to have a negative SARS-CoV-2 serologic test result at screening. Participants were excluded if they had a history of SARS-CoV-2 infection, a positive SARS-CoV-2 result at screening, or previous receipt of a vaccine or biologic agent indicated for the prevention of SARS-CoV-2 infection. All participants were randomized 1:1 to receive adintrevimab or placebo.

**Endpoint**

The primary objective for the PrEP cohort is to evaluate the efficacy of adintrevimab compared with placebo in the prevention of RT-PCR-confirmed symptomatic COVID-19 through 6 months, in participants with negative SARS-CoV-2 testing (RT-PCR and serology) at baseline as assessed by the proportion of participants with RT-PCR-confirmed symptomatic COVID-19 through 3 months. Secondary or exploratory endpoints include RT-PCR-confirmed symptomatic COVID-19 through 6 months, PK analyses, and safety analyses.

**Statistical analysis**

RT-PCR-confirmed symptomatic COVID-19 was determined by the protocol defined COVID-19 symptoms occurring within 14 days from the sample collection date of a positive central or local (in the absence of central test) RT-PCR. Any COVID-19-related hospitalization with a positive local SARS-CoV-2 test (within 14 days) or all-cause death are counted toward the endpoint. The date of event is the earliest date of COVID-19 symptom onset, COVID-19-related hospitalization, or all-cause death.

The analysis of the primary estimand, incidence of RT-PCR-confirmed symptomatic COVID-19 for adintrevimab versus placebo was analyzed using the methodology for determining a standardized estimator for a binary outcome with adjustment for the prognostic factors. The standardized risk difference, associated p-value and 95% CI, and the standardized relative risk reduction with 95% CI were provided for efficacy assessment.

A treatment policy strategy was used to handle the intercurrent events (ICEs) of interest in the primary analysis. The ICEs included the use of rescue medications (eg, COVID-19 vaccine, COVID-19 mAb for the purposes of prevention), subject unblinding by Investigator prior to the primary endpoint outcome, or others as specified in the SAP. Participants with missing primary endpoint outcome data were imputed as not having the primary endpoint outcome in the primary analysis.

**Viral testing, whole genome sequencing (WGS), and variants analysis**

Virological testing, WGS, and bioinformatics analysis of SARS-CoV-2 variants for the EVADE clinical study were performed at Eurofins Viracor BioPharma (Lenexa, Kansas, USA). A quantitative real-time RT-PCR (RT-qPCR) assay was used to detect SARS-CoV-2 infection from nasopharyngeal and/or saliva specimens during illness visits (*44*). For WGS, nucleic acid extraction for respiratory specimens was performed using the KingFisher (Thermofisher) with GSD NovaPrime® RNA Extraction (AE1) Kit (Eurofins). Extracted RNA was converted into cDNA using LunaScript™ Reverse Transcriptase (RT) Supermix (New England BioLabs). Following reverse transcription, cDNA was amplified using ARTIC SARS CoV2 Primer Pools (Eurofins Genomic Laboratories, EGL). PCR reactions from the pools were combined and subjected to magnetic bead clean up. The concentration of amplicons in each sample was quantified using the Qubit FLEX fluorometer (ThermoFisher) and Qubit 1X dsDNA HS reagents (ThermoFisher) and normalized. Preparation of libraries was performed using the NEBNext Ultra II FS library prep kit (New England Biolabs) in conjunction with the BRAVO liquid handling platform (Agilent). Following normalization, amplicons underwent enzymatic fragmentation and end repair using the NEBNext Ultra II FS library prep kit (New England Biolabs). Following end-repair fragmentation, AT-tailed uracil-linked hairpin adapters were ligated onto amplicons. After adapter ligation, USER enzyme was added to each reaction to cleave the uracil-linker in the hairpin of the adapter molecules. Following USER digest, automated purification of the library reactions was performed using the BRAVO liquid handler (Agilent) and SPRIselect magnetic beads (Beckman-Coulter). Index PCR Unique dual-indexed (UDI) primers (New England BioLabs) were used to barcode and amplify each individual library. Following index PCR, automated purification of the library reactions was performed using the BRAVO liquid handler (Agilent) and SPRIselect magnetic beads (Beckman-Coulter). The mass of each purified indexed library was quantified using the Qubit FLEX fluorometer (ThermoFisher) and Qubit 1X dsDNA HS reagents (ThermoFisher) and normalized. Libraries were pooled in equal volumes. The fragment size distribution of the final pooled library was confirmed using the TapeStation 4200 (Agilent) prior to preparation for sequencing. Pooled libraries were denatured, diluted, and sequenced on the Illumina NextSeq 500/550 instrument using a NextSeq Mid Output flow cell and reagents running a 2x150 cycle paired-end sequencing protocol.

Raw sequencing data (bcl files) were demultiplexed and converted to fastq files with bcl2fastq from Illumina. The fastq files were analyzed with the Eurofins Genomics SARS-CoV-2 NGS bioinformatic pipeline which trimmed primer sequences and filtered low quality reads, mapped high-quality reads to the reference genome (MN908947), called single nucleotide variants (snv) and small insertions & deletions (indel), generated a consensus sequence, and assigned SARS-CoV-2 lineages using pangolin (*45*) (cov-lineages.org)

**Pharmacokinetics of adintrevimab in NHPs**

A nonGLP NHP study was conducted at WuXi AppTec (Suzhou, China) to determine the serum pharmacokinetic (PK) properties of adintrevimab following a single intravenous (IV)

infusion or intramuscular injection (IM) administration. Twelve (12, 6/sex) naïve cynomolgus monkeys were divided into two groups with 3 animals/sex/group. Animals in the IV group were administered adintrevimab by a single 60-minute intravenous infusion administration at 10 mg/kg. Animals in the IM group were administrated adintrevimab by a single intramuscular injection at 10 mg/kg. Blood samples were collected at pre-dose (0), 1 (immediately after 1h infusion), 6, 24, 48, 96, 144, 168, 192, 336, 504, 672, 1008, 1344, 1680, 2016 and 2352 hours post-dose. Concentrations of adintrevimab in NHP serum samples were determined by a qualified ELISA method. The serum concentration-time profiles of adintrevimab in study animals were analyzed using a non-compartmental pharmacokinetic method (Phoenix

WinNonlin software (version 6.3, Pharsight, Mountain View, CA)).

**Bioanalytical assay to measure adintrevimab concentration in NHPs**

Quantification of adintrevimab in NHP serum was performed using a qualified bioanalytical immunoassay method at WuXi AppTec (Suzhou, China). SARS-CoV-2 Spike protein (RBD from GenScript) was coated onto a microplate, and NHP serum samples, QCs, or standards were added to coated wells. Following incubation, serum was removed, wells were washed, and mouse anti-human IgG Fc-HRP detection antibody was added. TMB substrate (SeraCareTM) including low pH stop, was used for detection, followed by absorbance reading at 450 nm/630 nm. Concentrations of adintrevimab in samples and QCs were determined by back-calculation to a standard curve.

**Bioanalytical assay to measure adintrevimab concentration in human serum**

Quantification of adintrevimab in human serum was performed using a validated bioanalytical method at Q2 Solutions (Ithaca, NY). Adintrevimab was isolated from serum-based calibration standards, quality control samples (QC) and subject samples via immunoprecipitation in which samples were combined with biotinylated monoclonal anti-adintrevimab (anti-idiotypic antibody) which specifically binds adintrevimab in samples. Streptavidin beads were used to capture the biotinylated antibody-adintrevimab complex. Adintrevimab was then digested with trypsin to yield signature peptides specific to adintrevimab. A stable isotope-labeled signature peptide internal standard was then added to the calibration standards, QCs, and subject samples. All samples were assayed using liquid chromatography/tandem high-resolution mass spectrometry (LC/HRMS/MS). All concentration calculations are based on the peak area ratio (PAR) of adintrevimab signature peptide to the internal standard. Concentrations of the analyte in QC and subject samples were determined by back-calculation from the calibration curve.

**Phase 1 clinical study serum neutralization assay**

Clinical samples were evaluated for serum neutralizing antibodies to SARS-CoV-2 using a microneutralization assay at Viroclinics Biosciences (Rotterdam, The Netherlands). The source of SARS-CoV-2 isolates included Delta: hCoV-19/USA/MD-HP05647/2021 (BEI Resources NR-55672), and Omicron BA.1: hCoV-19/Netherlands/NH-RIVM-72291/2021 (European Virus Archive). Briefly, virus was preincubated with serial dilutions of serum, then the virus-serum mixtures were added to a confluent monolayer of Vero-E6 cells. After 1 hour of incubation, cell media was replaced, and cells were incubated for an additional 15-23 hours. Cells were then fixed with formalin, followed by immunostaining using a primary antibody targeting the viral nucleocapsid protein and a peroxidase-conjugated secondary reagent. Virus positive cells were detected using TrueBlue substrate and quantified using an ImmunoSpot analyzer (Cellular Technology Limited). The 50% neutralization titers (i.e. MN50) were calculated according to the method described by Zielinska et al. (*46*).

**Population PK model development**

A population pharmacokinetic (PK) model was developed for ADG20 using serum ADG20 concentration-time data from the two ongoing clinical trials as well as a Phase 1, first-in-human study (ADG20-1-001). The development of the population PK model involved three steps: 1) construction of the base structural/statistical model, 2) conduct of a covariate analysis to identify subject descriptors associated with the interindividual variability (IIV) in PK, and 3) evaluation and qualification of the final model. Base structural model development was initially conducted using the Phase 1, intensive PK sampling data only and consisted of the fitting of 1, 2, and 3 compartmental population PK models with linear elimination and first-order drug absorption. The base structural model was then fit to the pooled Phase 1 and Phase 2/3 data and refined as necessary to assure a robust fit across all subjects. The potential for target-mediated drug disposition was evaluated as necessary based upon comparisons of the observed data in infected subjects relative to those that were not infected. After an appropriate base structural model was identified, population PK covariate model development was undertaken using forward selection followed by a backward elimination procedure. The resultant final population PK model was then evaluated for potential revisions such as the removal of extraneous covariate relationships or modifications to the interindividual and residual variability models. The model was then qualified by performing a prediction-corrected visual predictive check (PC-VPC), which graphically examines the agreement between the 5th, 50th, and 95th percentiles of the observed and the individual simulated concentrations across time intervals. The population PK analysis was conducted using NONMEM^®^ software Version 7.4 (ICON Development Solutions, Ellicott City, MD) implementing the first-order conditional estimation method with interaction [reference]. Model-based simulations were conducted by converting the NONMEM model code to C++ code so that simulations could be conducted using mrgsolve, a package for R statistical software that facilitates simulations from differential equation-based models.

**Fc**g**R binding studies**

SPR analysis was conducted at 25 °C in a HBS-EP+ buffer system (10 mM HEPES, pH 7.4, 150 mM NaCl, 3 mM EDTA, 0.05% Surfactant P20) using a Biacore 8K optical biosensor equipped with a CAP sensor chip (Global Life Sciences Solutions USA, Marlborough, MA). (Global Life Sciences Solutions USA, Marlborough, MA). The sample compartment was maintained at 10°C for the duration of the experiment. This assay orientation allows for reproducible capture of biotinylated samples to the sensor surface. Prior to each analysis the sensor chip surface was first conditioned with 3 pulses (60 s at 10 ml/min) of regeneration solution (6 M Guanidine-HCl in 0.25 M NaOH). Each experiment cycle began with an injection (300 s at 2 ml//min) over flow cells 1 and 2 of a 1:20 solution of biotin CAPture reagent (Global Life Sciences Solutions USA, Cat.# 29423383) in HBS-EP+ buffer. This was followed by an injection (180 s at 10 ml/min) of the biotinylated RBD antigen over flow cell 2. Upon capture of the antigen to the sensor surface, ADG2 or adintrevimab was injected (180 s at 30 ml/min) over flow cells 1 and 2. The dissociation of the IgG was monitored for 120 s prior to injection (180 s) of the FcgR. Dissociation of the FcgR from the sensor surface was monitored for 180 s. Finally, an injection (120 s at 10 ml/min) of regeneration solution over flow cells 1 and 2 prepares the sensor surface for another cycle. The data was first cropped to include only the steps that involve the FcgR association and dissociation. This selected data was then aligned, double reference subtracted, and then non-linear least squares fit to a 1:1 binding model using Biacore Insight Evaluation software version 3.0.11.15423.

**FcRn binding studies**

SPR analysis was conducted at 25 °C using a Biacore 8K optical biosensor equipped with either a CM3 or CM5 sensor chip (Global Life Sciences Solutions USA, Marlborough, MA). The sample compartment was maintained at 10°C for the duration of the experiment. These studies were conducted in an HBS-EP+ buffer system (10 mM HEPES, 150 mM NaCl, 3 mM EDTA, 0.05% Surfactant P20) at pH 6.0 or pH 7.4. A pH scouting study helped determine the buffer pH and approximate concentration for direct immobilization of each IgG to the sensor surface. The sensor surface was prepared as follows: a 1:1 mixture of EDC and NHS was injected (420 s) over flow cells 1 and 2, the antibody was injected (120 s) over flow cell 2 and finally ethanolamine was injected (420 s) over flow cells 1 and 2. Each experiment cycle began with an injection (180 s at 30 ml/min) of FcRn over flow cells 1 and 2. The dissociation of the FcRn was observed for 180 s before the sensor surface was regenerated via two injections (20 s at 30 ml/min) of HBS-EP+ buffer (pH 7.4) which prepares the sensor surface for another cycle. The data was aligned, double reference subtracted, and then non-linear least squares fit to a 1:1 binding model using Biacore Insight Evaluation software version 3.0.11.15423.

**C1q binding studies**

BLI analysis was conducted at 25 °C in a PBSF buffer system (phosphate buffered saline, pH 7.4, with 0.1% BSA) using a ForteBio Octet HTX (Sartorius Bioanalytical Instruments, Bohemia, NY) equipped with SA sensor tips. These assay conditions were adapted from Zhou et al (*47*). The sensor tips were soaked in PBSF buffer for 10 minutes and then exposed (60 s) to wells containing biotinylated RBD antigen. Each experiment cycle began with dipping (180 s) the sensor tip into PBSF to establish a stable baseline. This was followed by exposure (180s) of the antigen loaded sensor tip to wells containing the IgG. After a short dip (60 s) into fresh wells of PBSF, the sensor tip was dipped (180 s) into wells containing the C1q or blank buffer. The sensor tips were then immediately dipped (180 s) into fresh wells containing PBSF buffer to monitor (first 30 s) the dissociation of C1q from the sensor tip surface. The data was x and y-axis aligned and then non-linear least squares fit to a 1:1 binding model using ForteBio Data Analysis software version 11.1.3.10.

**Antibody-dependent natural killer cell activation and degranulation (ADNKDA)**

Primary human NK cells were enriched from the peripheral blood of human donors using RosetteSep Human NK cell Enrichment Cocktail (Stem Cell Technologies, Cat #15065) and cultured overnight in RPMI-1640 (Corning, Cat # 15-040-CV) supplemented with 10% FBS (Hyclone, Cat # SH30071.03), 1% Pen/Strep (Gibco, Cat # 15070-063), 1% L-Glutamine (Corning, Cat # 25-005-CI), 1% HEPES (Corning, Cat # 25-060-CI) and 5 ng/ml recombinant human IL-15 (StemCell Technologies, Cat # 78031). Recombinant SARS-CoV-2 receptor binding domain was coated onto MaxiSorp 96-well plates (Thermo Scientific, Cat # 442404) at 200 ng/well at 4 °C overnight. Wells were washed with PBS and blocked with 5% BSA prior to addition of antibodies that were diluted in a five-fold dilution series in PBS (10 µg/ml - 0.32 ng/ml) and incubation for 2 h at 37 °C. Unbound antibodies were removed by washing with PBS were added at 5 x 10^4^ cells/well in the presence of 4 µg/ml brefeldin A (Biolegend, Cat # 420601), 5 µg/ml GolgiStop (BD Biosciences, Cat # 554724) and anti-CD107a antibody (Clone H4A3 PE-Cy7, Biolegend, Cat # 328618) for 5 hours. Cells were stained for surface expression of CD16 (Clone 3G8 Pacific Blue, Biolegend, Cat # 302032), CD56 (clone 5.1H11 AlexaFluor488, Biolegend, Cat # 362518) and CD3 (clone UCHT1 Alexa Fluor700, Biolegend, Cat # 300424). Cells were fixed and permeabilized with Fix/Perm (Biolegend, Cat # 421002) according to the manufacturer’s instructions to stain for intracellular IFNγ (Clone B27 PE, Biolegend, Cat # 506507) and TNFα (clone Mab11 APC, Biolegend, Cat # 502912). Cells were analyzed on a Cytek Aurora spectral flow cytometer.

**Antibody-dependent cellular phagocytosis (ADCP) with monocytes and neutrophils**

For ADCP assays with neutrophils, HL-60 promyeloblast cells (ATCC, Cat # CCL-240) were maintained in Iscove's Modified Dulbecco's Medium (ATCC, Cat # 30-2005) with 20% fetal bovine serum and 1% Pen/Strep. HL-60 cells were differentiated into neutrophils by growth for 5 days in the presence of 1.3% DMSO. Recombinant SARS-CoV-2 RBD protein was coupled to fluorescent beads (Thermo Scientific, Cat # F8819) by carbodiimide coupling. Antibodies were diluted in a five-fold dilution curve in HL-60 culture medium (1000 - 0.32 ng/ml) and incubated with RBD-coated beads for 2 hours at 37 °C. Cells (5 x 10^4^/well) were incubated for 18 hours at 37 °C. Cells were then stained for CD11b (Clone M1/70 APC-Fire750, Biolegend, Cat # 101262) and CD16 (Clone 3G8 Pacific Blue, Biolegend, Cat # 302032), fixed with 4% paraformaldehyde, and analyzed by flow cytometry. CD11b+ and CD16+ cells were analyzed for uptake of fluorescent beads. A phagocytic score was determined using the following formula: (percentage of FITC^+^ cells)*(geometric mean fluorescent intensity (gMFI) of the FITC^+^ cells)/100,000.

For ADCP assays with monocytes, THP-1 monocytes were maintained in RPMI-1640 supplemented with 10% FBS, 1% Pen/Strep, 1% L-glutamine, and β-mercaptoethanol. Recombinant SARS-CoV-2 RBD-coated beads were generated as described above. Antibodies were diluted in a five-fold dilution curve in THP-1 culture medium to (5000–0.064 ng/ml) and incubated with RBD-coated beads for 2 h at 37 °C. Unbound antibodies were removed by centrifugation prior to the addition of THP-1 cells at 2.5 x 10^4^ cells/well. Cells were fixed with 4% paraformaldehyde and analyzed by flow cytometry. A phagocytic score was determined as described above.

**Antibody-mediated complement deposition (ADCD)**

Recombinant SARS-CoV-2 receptor binding domain-coated beads were generated as described for ADCP assays. Antibodies were diluted in a five-fold dilution series in RPMI-1640 (5000 - 0.064 ng/ml) and incubated with RBD-coated beads for 2 hours at 37 °C. Unbound antibodies were removed by centrifugation prior to the addition of reconstituted guinea pig complement (Cedarlane Labs, Cat # CL4051) and diluted in veronal buffer supplemented with calcium and magnesium (Boston Bioproducts, Cat # IBB-300) for 20 minutes at 37 °C. Beads were washed with PBS containing 15 mM EDTA, and stained with an FITC-conjugated anti-guinea pig C3 antibody (MP Biomedicals, Cat # 855385). C3 deposition onto beads was analyzed by flow cytometry. The gMFI of FITC for all beads was measured.

**Polyreactivity assay**

Polyspecificity reagent binding of antibodies was performed as described previously (*19*). Briefly, soluble membrane protein (SMP) and soluble cytosolic protein (SCP) fractions were extracted from Chinese hamster ovary (CHO) cells and biotinylated using NHS-LC-Biotin (Thermo Fisher Scientific) reagent. Yeast-presented IgGs were incubated with 1:10 diluted stock of biotinylated SMP and SCP for 20 minutes on ice, followed by two washes with PBSF, and stained with 50 µL of a secondary labeling mix containing ExtrAvidin-R-PE (Sigma-Aldrich), anti-human LC-FITC (Southern Biotech), and propidium iodide (Invitrogen) for 15 minutes on ice. Cells were subsequently washed with PBSF and resuspended in PBSF for flow cytometric analysis on a BD FACS Canto II (BD Biosciences).

**Affinity-capture self-interaction nanoparticle spectroscopy (AC-SINS)**

To measure the propensity for antibodies to self-associate, AC-SINS was performed as previously described (*19*). Briefly, polyclonal goat anti-human IgG Fc antibodies (capture; Jackson ImmunoResearch Laboratories) and polyclonal goat non-specific antibodies (non-capture; Jackson ImmunoResearch Laboratories) were buffer exchanged into 20 mM sodium acetate (pH 4.3) and concentrated to 0.4 mg/ml. A 4:1 volume ratio of capture:non-capture was prepared and further incubated at a 1:9 volume ratio with 20 nm gold nanoparticles (AuNP; Ted Pella Inc.) for 1 hour at room temperature (RT). Thiolated PEG (Sigma-Aldrich) was then used to block empty sites on the AuNP and filtered via a 0.22 μm PVDF membrane (Millipore). Coated particles were subsequently added to the test antibody solution and incubated for 2 hours at RT before measuring absorbance from 510 to 570 nm on a plate reader. Data points were fit with a second-order polynomial in Excel to obtain wavelengths at maximum absorbance. Values are reported as the difference between plasmon wavelengths of the sample and background (Δλ_max_).

**Fab thermal stability**

Apparent melting temperatures (T_m_^App^) of Fab fragments were obtained as previously described (*19*). Briefly, 20 μl of test antibody solution at 1 mg/ml was mixed with 10 μl of 20 × SYPRO orange. The plate was scanned with a CFX96 Real-Time System (BioRad) from 40 °C to 95 °C at a rate of 0.25 °C/minute. T_m_^App^ was calculated from the primary derivative of the raw data via the BioRad analysis software.

**Hydrophobic interaction chromatography (HIC)**

Antibody hydrophobicity was evaluated using HIC as previously described (*19*). Briefly, test antibody samples were diluted in phase A solution (1.8 M ammonium sulfate and 0.1 M pH 6.5 sodium phosphate) to a final concentration of 1.0 M ammonium sulfate. A linear gradient from phase A solution to phase B solution (0.1 M pH 6.5 sodium phosphate) was run for 20 minutes at a flow rate of 1.0 ml/minute using the Sepax Proteomix HIC butyl-NP5 column. Peak retention times were obtained from monitoring UV absorbance at 280 nm.

Supplementary Figures

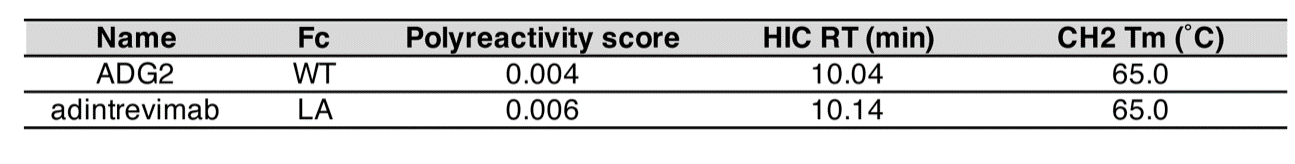

Fig. S1. Biophysical properties of ADG2 and adintrevimab. (A) Antibody polyreactivity score, as assessed based on binding to a previously described polyspecificity reagent (*19*). (B) Antibody hydrophobicity, as evaluated by hydrophobic interaction chromatography (*19*). (C) IgG CH2 thermal stability, as determined by differential scanning fluorimetry (DSF). RT; retention time.

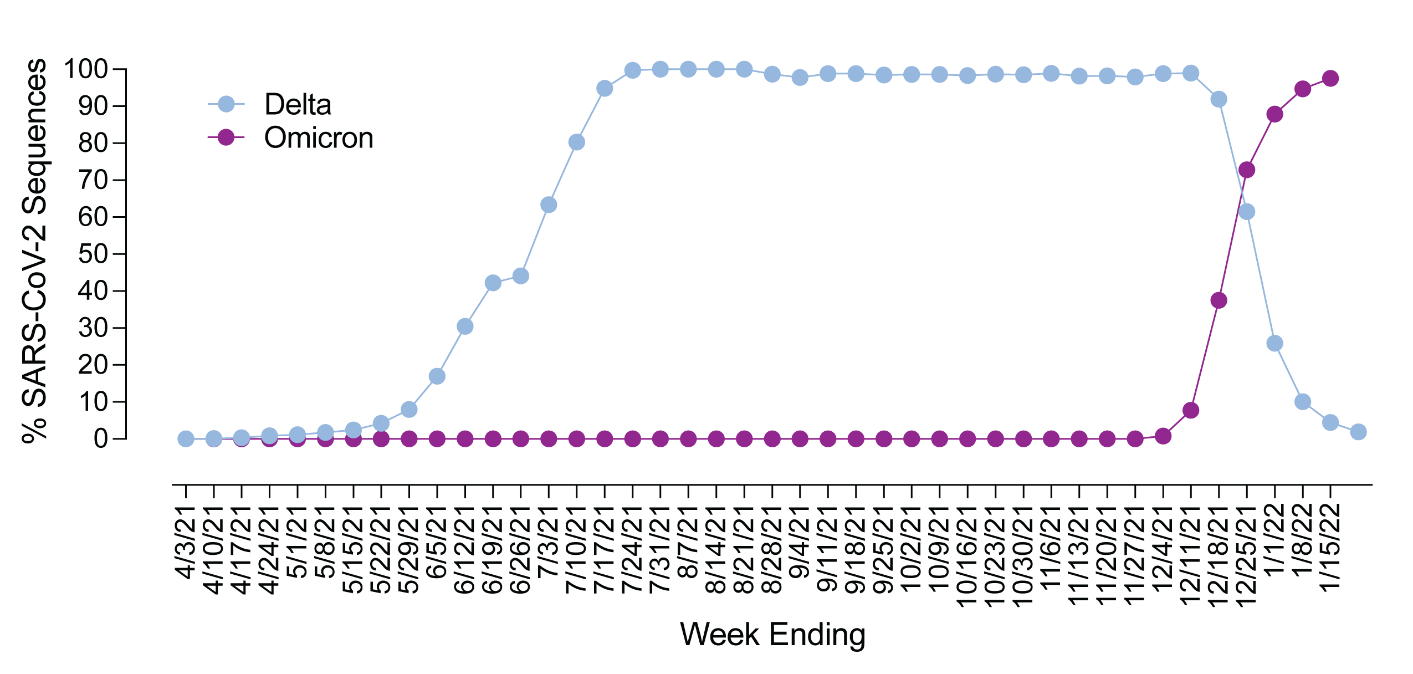

Fig. S2. SARS-CoV-2 variant frequency in surveillance sequencing. Weekly estimated frequencies of SARS-CoV-2 Delta and Omicron BA.1/BA1.1 variants between weeks ending on April 4, 2021 and January 15, 2022 in the United States were retrieved from a publicly available dataset obtained through the CDC National SARS-CoV-2 Strain Surveillance (NS3) system (*48*). Cumulative Delta and Omicron BA.1/BA1.1 frequencies represent the combined frequencies of Pango lineages B.1.617.2, AY.1, AY.2, and AY.3; and B.1.1.529 (BA.1) and BA.1.1 lineages, respectively. Potentially unreliable data from weeks where estimates totaled less than 10 sequences were excluded from the analysis.

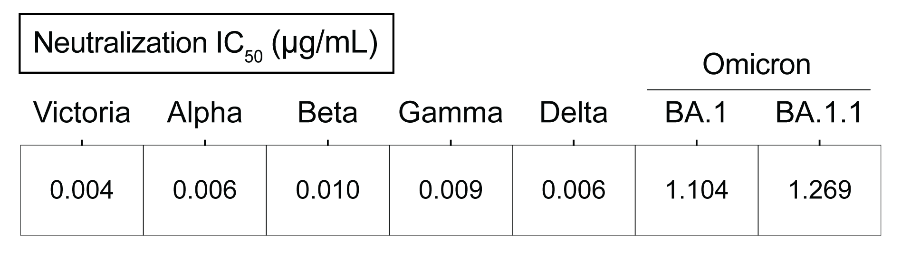

**Fig. S3**. Neutralization of SARS-CoV-2 variants by adintrevimab. Neutralization IC_50_s were determined using a focus reduction neutralization test (FRNT) and previously reported by Dejnirattisai et al. (*11, 21, 22*).

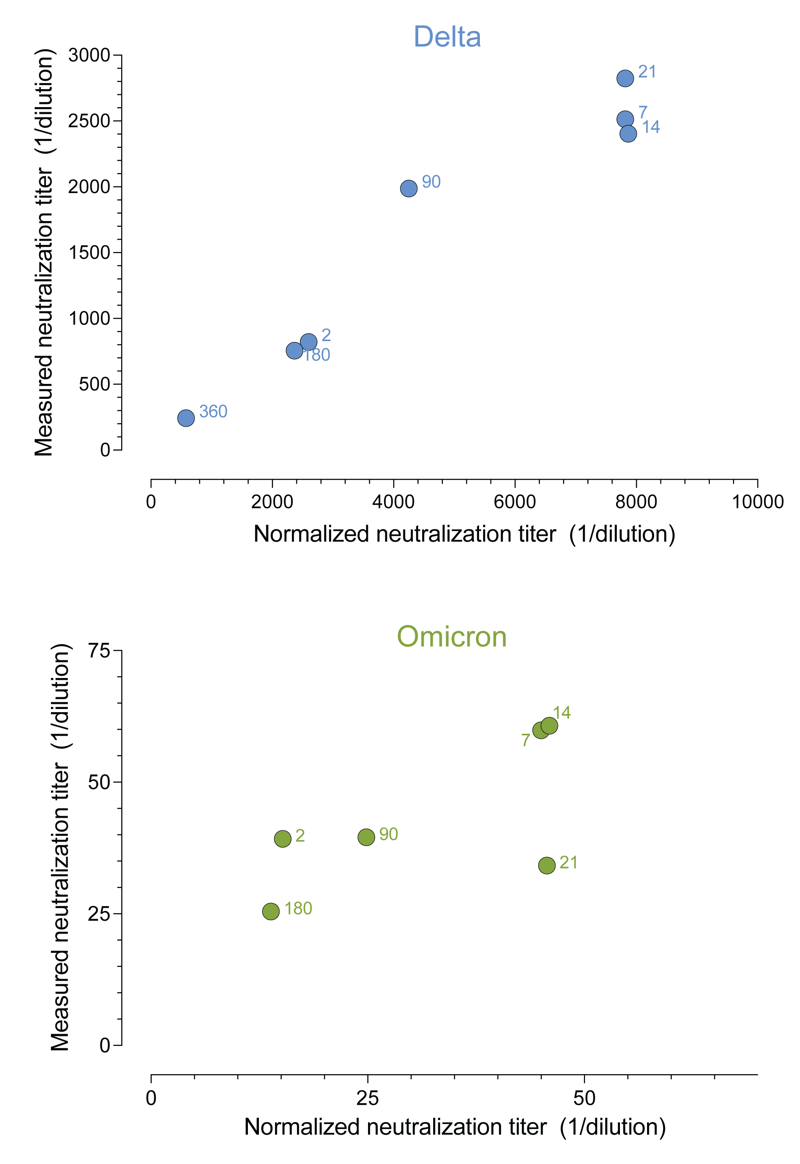

Fig. S4. Correlation between experimental and normalized serum neutralizing titers for Delta and Omicron BA.1. Normalized serum neutralizing titers were generated by dividing population PK model median adintrevimab serum concentrations by experimentally determined authentic virus neutralization IC_50_s for adintrevimab against Delta and Omicron BA.1/BA1.1 (serum neutralization titer = adintrevimab serum concentration/variant IC_50_) (*11, 21, 22*). Experimental serum neutralizing titers were determined in an authentic virus FRNT assay using serum samples from participants enrolled in our Phase 1 study. Numbers shown next to data points indicate the number of days following adintrevimab dosing that serum samples were collected.

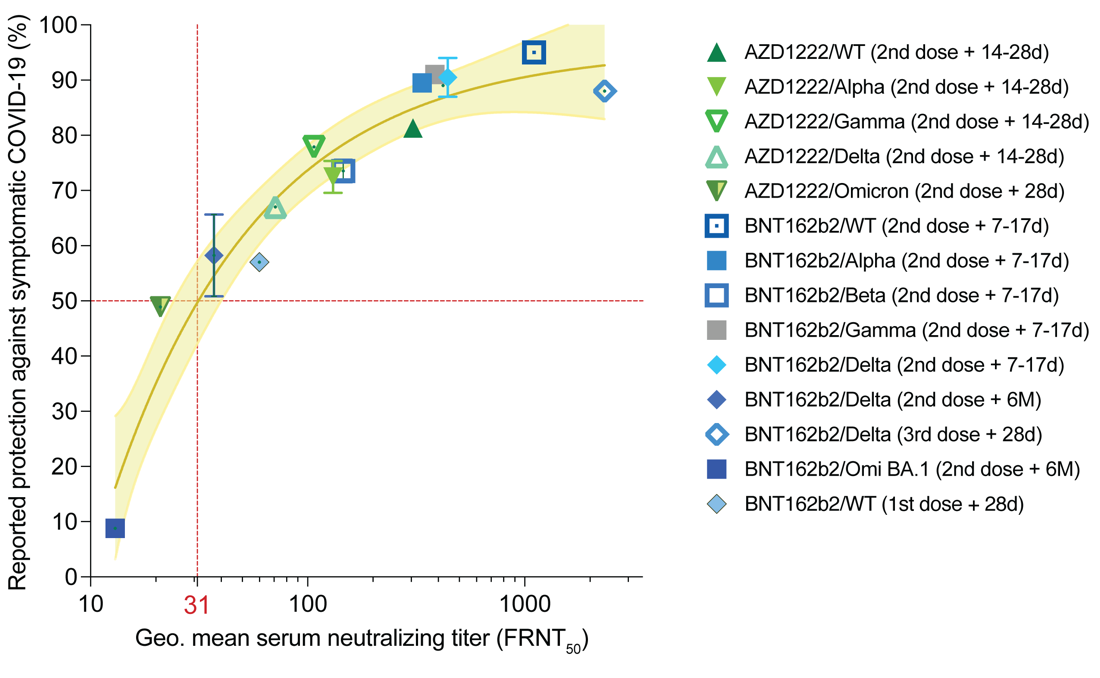

**Fig. S5.** Relationship between median serum neutralization titer and protective efficacy against symptomatic COVID-19. (A) Reported median vaccine-induced serum neutralizing titer, measured in an authentic virus neutralization assay (*11, 21, 22*), plotted against reported efficacy in Ph3 clinical trials and/or real-word vaccine effectiveness studies (*23-37*). The number of vaccine doses and timing of blood sample collection and corresponding vaccine effectiveness study read-outs against the indicated variants are shown in parenthesis in the legend. The brown solid line indicates the best fit of the non-linear regression and the yellow shading indicates 95% confidence intervals. Data points and error bars represent mean ± SD. The neutralizing titer associated with 50% protection against symptomatic COVID-19 is indicated by a vertical dotted red line. “WT” refers to ancestral SARS-CoV-2 strains (Wuhan-1 and D614G). Omi, Omicron; Geo., geometric.

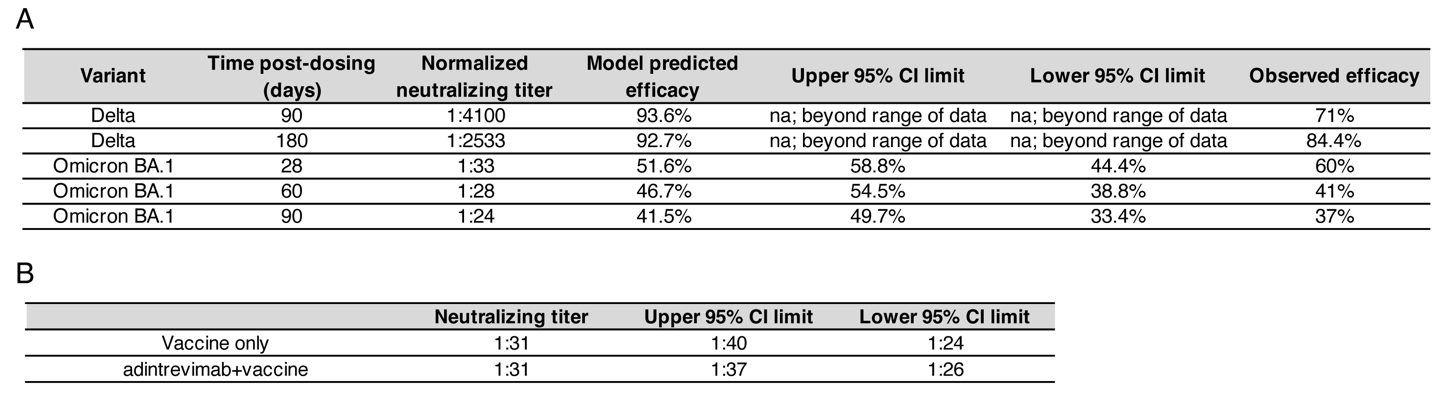

**Figure S6.** (A) Observed and model predicted efficacy of adintrevimab against symptomatic Delta and Omicron BA.1/BA1.1 infection. Predicted model efficacy and 95% confidence intervals were determined via non-linear regression analysis of protection versus serum neutralizing titers (as shown in Figure S5) and statistical interpolation of efficacy from normalized neutralizing titers at the indicated days post-dosing. Confidence intervals could not be calculated for extrapolated efficacies against Delta because the neutralizing titers were higher than the maximum neutralizing titers achieved by vaccination. CI = confidence interval.

(B) Predicted neutralization titer associated with 50% protection against symptomatic COVID-19 based on vaccine data (top row) and vaccine and adintrevimab data (bottom row). Serum neutralizing titers associated with 50% protection were interpolated from non-linear regression analyses of the correlation between efficacy and serum neutralizing titers (curves shown in Fig. 4 and Fig. S5). CI = confidence interval.

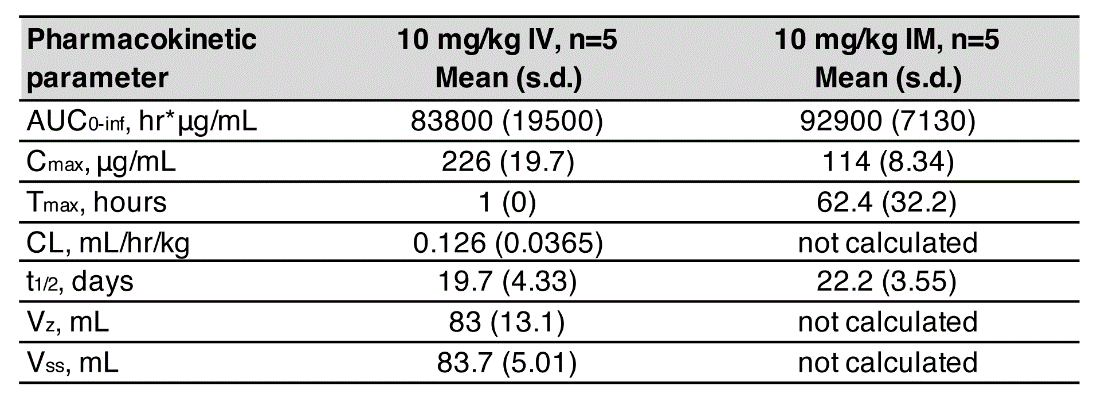

**Table S1.** Adintrevimab pharmacokinetic parameters in NHPs.

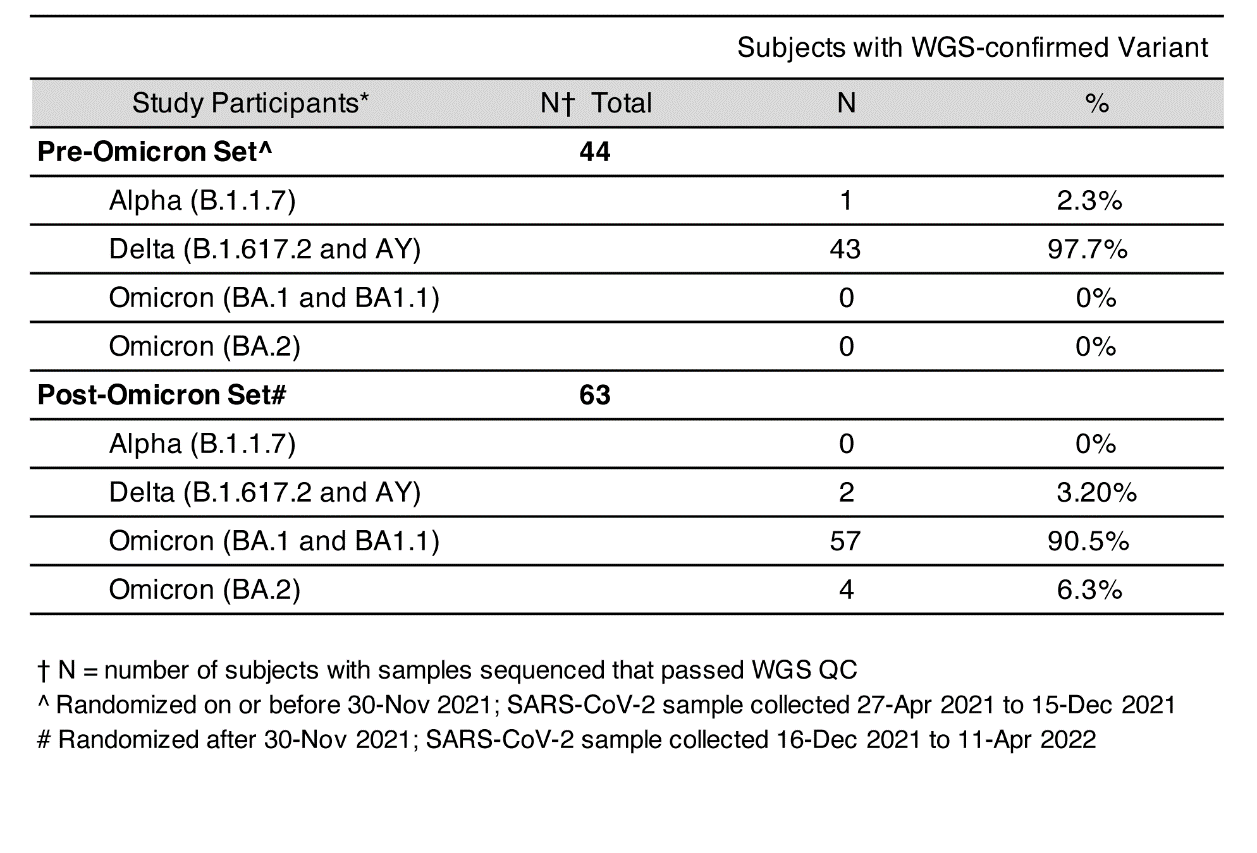

**Table S2.** Summary of SARS-CoV-2 variants observed in EVADE study analysis sets.

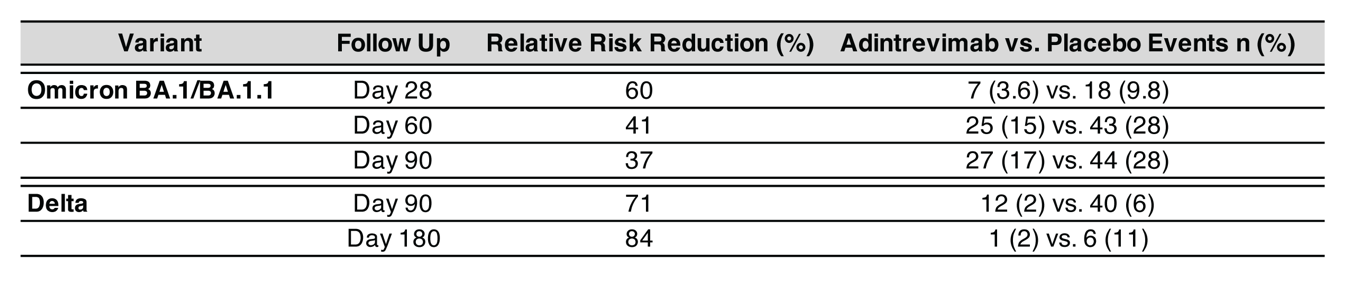

**Table S3.** Primary end point and additional supportive efficacy analyses from EVADE.

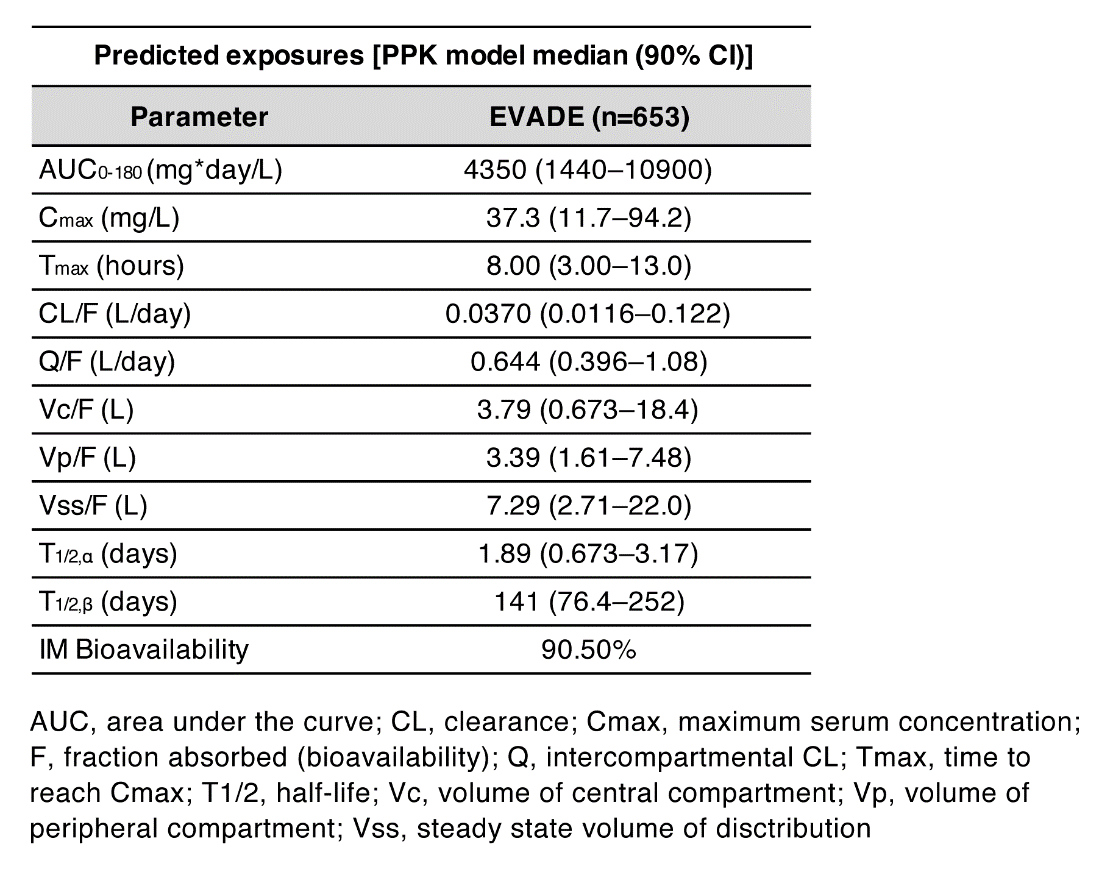

**Table S4.** Population pharmacokinetic parameters for a 300 mg IM dose of adintrevimab in humans.

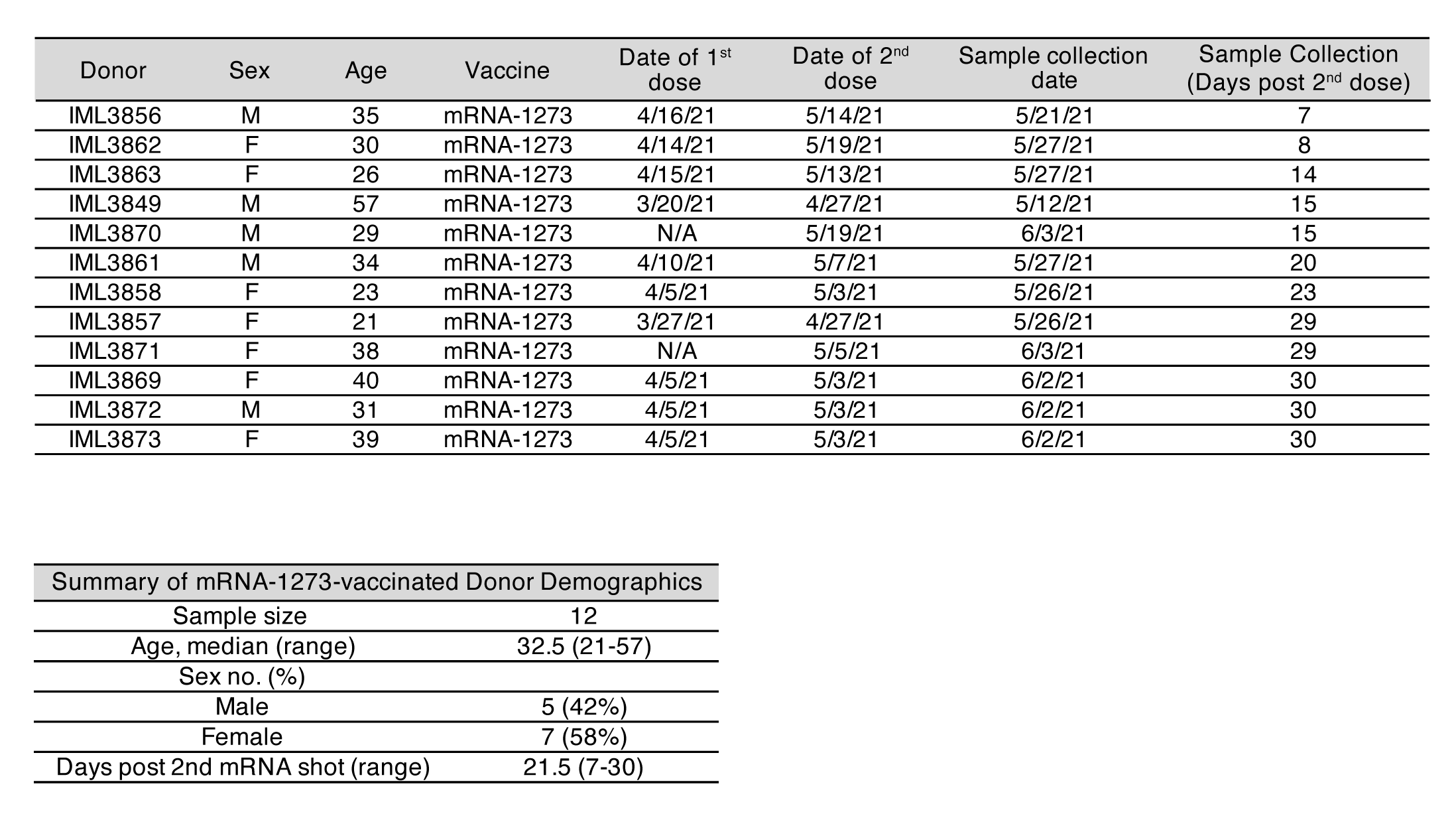

**Table S5.** mRNA-1273 vaccinated donor information.

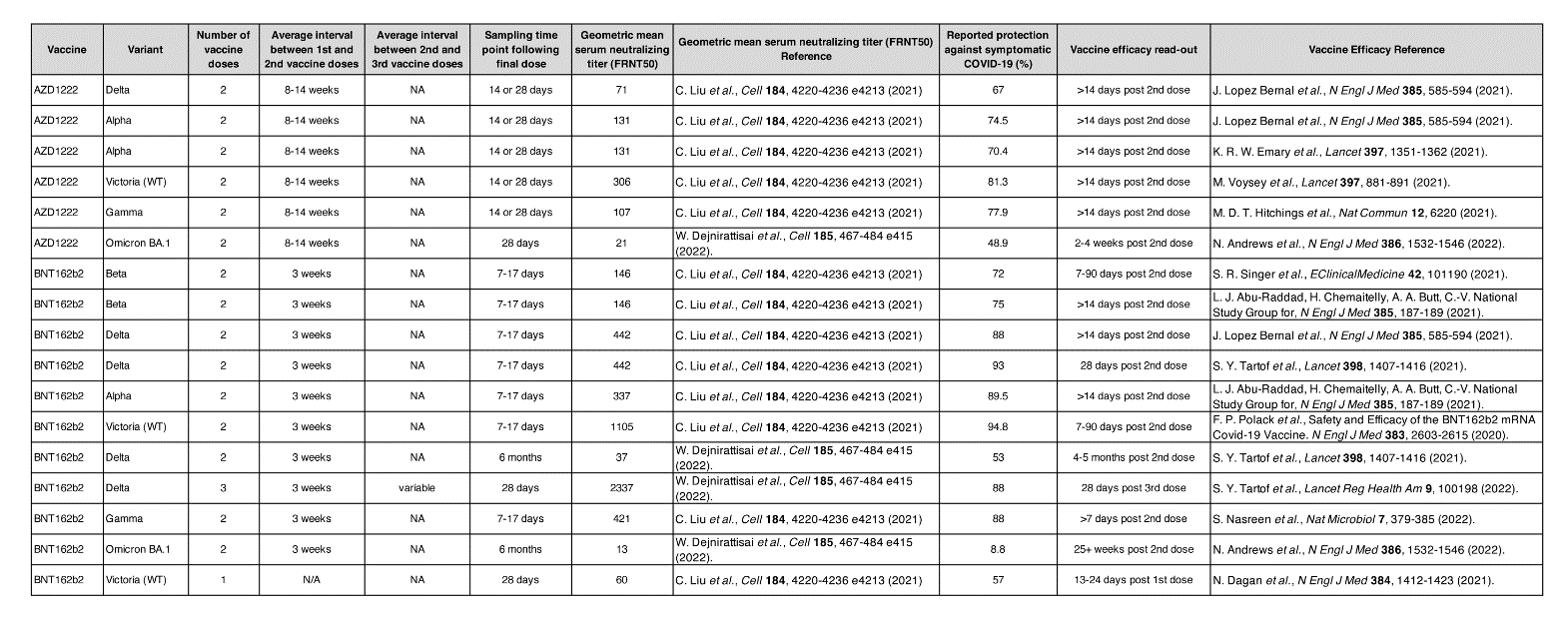
**Table S6.** Data sources for vaccine efficacy and immunogenicity analyses.
